# Bi-allelic *NIT1* variants cause small vessel disease with movement disorders and massive non-lobar intracerebral haemorrhage

**DOI:** 10.1101/2023.05.16.23289676

**Authors:** J.W. Rutten, M.N. Cerfontaine, K.L. Dijkstra, A.A. Mulder, M. Kruit, R. de Koning, S.T. de Bot, K.M. van Nieuwenhuizen, J.J. Baelde, H.W. Berendse, G.J.G Ruijter, F. Baas, C.R. Jost, S.G. van Duinen, E.A.R. Nibbeling, G. Gravesteijn, S.A.J. Lesnik Oberstein

## Abstract

Highly penetrant monogenic causes of intracerebral haemorrhage are rare, and are almost exclusively hereditary cerebral amyloid angiopathies caused by heterozygous pathogenic variants in the *APP* gene. Here, we identified a novel genetic cause of mid-adult onset non-lobar ICH, caused by bi-allelic pathogenic variants in the *NIT1* gene. The seven identified patients from five unrelated pedigrees presented with movement disorders, slowly progressive cognitive decline, ischemic strokes and psychiatric disturbances. All patients shared a striking neuroimaging phenotype with a honeycomb appearance of the basal ganglia due to an extremely high burden of enlarged perivascular spaces. Two patients were deceased, due to mid-adult massive non-lobar intracerebral haemorrhage. Small cerebral arteries showed strongly abnormal morphology, with thickening of the media and numerous large electron dense deposits located between the media and adventitia. Patients were homozygous for the *NIT1* c.727C>T; p.Arg243Trp variant or compound heterozygous for the *NIT1* c.727C>T; p.Arg243Trp and c.198_199del, p.Ala68* variant. Urine analysis showed increased levels of deaminated gluthatione, consistent with loss of NIT1 function in both homozygous and compound heterozygous patients. Based on *NIT1* carrier frequencies in UK Biobank and gnomAD, an estimated minimum of 4500 individuals worldwide are affected with this novel, autosomal recessively inherited cause of intracerebral haemorrhage, which we term NIT1-small vessel disease.

## Introduction

Intracerebral haemorrhage (ICH) is caused by small vessel disease (SVD) in up to 85% of patients.^1^ The main cause of lobar ICH is cerebral amyloid angiopathy, whereas non-lobar ICH is mostly attributable to hypertensive angiopathy.^2^ In a minority of individuals with ICH, there is a positive family history of early onset ICH. Highly penetrant monogenic causes of ICH are very rare. Genetic disorders in which ICH is the cardinal feature are almost exclusively caused by heterozygous pathogenic variants in the *APP* gene, leading to diverse subtypes of hereditary cerebral amyloid angiopathy characterized by mid-adult onset lobar ICH.^3^ *NOTCH3, COL4A1* and *COL4A2* are other examples of genetic small vessel disease (SVD) in which ICH can occur, but with a much lower frequency.^4^ Autosomal recessive forms of ICH have to the best of our knowledge never been reported.

Here, we identified a novel, autosomal recessive metabolic SVD with mid-adult onset non-lobar ICH preceded by various movement disorders, and a highly distinct neuroimaging phenotype with massively enlarged perivascular spaces in the basal ganglia-thalamus complex, caused by bi-allelic *NIT1* variants. We describe the clinical, neuroimaging, pathological, genetic and biochemical profiles of seven affected individuals from five unrelated Dutch pedigrees and calculated minimal estimated disease prevalence based on *NIT1* pathogenic variant frequencies from the GoNL database, gnomAD and UK Biobank.

## Materials and methods

### Subjects

The first ascertained family comprised three siblings with a distinct neuroimaging phenotype and ICH, leading to the suspicion of an autosomal recessive monogenic cause. Subsequently, two unrelated and isolated cases with a similar neuroimaging phenotype were recognized and two unrelated and isolated cases were identified after re-analysis of next generation sequencing data in patients previously referred to the Leiden Laboratory for Diagnostic Genome Analysis because of suspected genetic small vessel disease. This study has been approved by the Medical Ethics Committee Leiden The Hague Delft (P21.013). Written informed consent was obtained from all patients.

### Exome sequencing and 3D modelling

Genomic DNA was extracted from whole blood. Exome sequencing was performed on a Illumina platform after exome enrichment using the Agilent SureSelectXT Human All Exon v5,v7 or SureSelect Clinical Research Exome V2 kit at Genomescan B.V., Leiden, the Netherlands. Burrows-Wheeler Aligner was used for read alignment and Genome Analysis Tool Kit was used for variant calling. Annotation was performed using in-house LUMC software using reference transcript NM_005600.2. 3D modelling was performed using AlphaFold and 3D models were visualized using ChimeraX.

### Neuroimaging

For the two affected patients who participated in our on-site clinical study, brain MRI was performed on a 3 Tesla MR system (Philips Achieva TX, Philips Medical Systems), including the following sequences: 3-dimensional T1-weighted images, T2-weighted images, fluid-attenuated inversion recovery and susceptibility-weighted images. For all other patients, either clinical 1.5 tesla MRIs were available with exception of one patient, who had only undergone a CT-scan at the time of a massive haemorrhagic stroke. Brain MRIs were qualitatively assessed by MC and by MK, an experienced neuroradiologist specialized in small vessel disease.

### Brain and skin vessel immunohistochemistry and electron microscopy

Paraffin embedded brain tissue was available of two deceased siblings, frozen brain tissue was available of one. Control brain tissue was obtained from two deceased individuals in whom autopsy was performed and tissue processed at the same pathology department as that of the patients. Brain tissues was sectioned and stained with Haematoxylin-eosin, Periodic acid-Schiff, toloidine blue, and Congo Red. Immunohistochemistry was performed using NOTCH3 (1E4, Millipore, dilution 1:1000), NIT1 (Ab180942, Abcam, dilution 1:50) and beta amyloid antibodies (Clone 6F/D3, Dako, dilution 1:20). Skin biopsies were obtained from three patients and three unaffected heterozygous family members, and were processed for immunohistochemistry and electron microscopy. Skin tissue was also snap frozen and stored for fibroblast culture. Electron microscopy was performed in brain tissue of the two deceased patients and in two skin biopsies.

### Metabolic analysis

Urine samples were obtained from two NIT1-SVD patients and three unaffected heterozygous family members. Analysis of urine organic acids, including the NIT1 substrate deaminated gluthatione (dGSH),^5^ was performed by gas chromatography-mass spectrometry of trimethylsilyl-derivatives.

### Ascertainment of *NIT1* variants in UK Biobank

Details on the UK Biobank study have been described previously.^6^ *NIT1* truncating variants (nonsense and frameshift variants) and the *NIT1* c.727C>T; p.Arg243Trp variant were ascertained in 454 805 whole-exome data using reference transcript NM_005600.2. Of individuals with these *NIT1* variants, ICD10 codes and available brain MRI’s were extracted. Brain MRI was available for 20 c.727C>T; p.Arg243Trp carriers and for 42 carriers of a truncating *NIT1* variant, and were compared to 62 age and sex matched controls from UKB.

### Analysis of stroke frequency and MRI quantification in UK Biobank *NIT1* variant carriers

Ischemic and hemorrhagic stroke were assessed using ICD-10 codes (I61, I63, I64). The following conventional SVD markers were quantitively evaluated according to consensus criteria^7^: normalized WMH volume (nWMHv) and burden of enlarged perivascular spaces (ePVS). All MRIs from UKB were rated by an experienced observer (MC) blinded for genotype. WMH were quantified using the Brain Intensity Classification Algorithm.^8^ and normalized to the intracranial volume (normalized WMH volume=[total WMH volume/intracranial volume]×100). ePVS and lacunes were evaluated on T_1_-weighted images. ePVS were evaluated in the following regions: basal ganglia, subinsular region, global white matter and temporal lobe according to a previously described 4-grade semiquantitative scale.^9^ In addition, due to its extensive presence in NIT1-SVD, ePVS were also scored in the occipital lobe, cerebellar and in the mesencephalon.

## Results

### Genetics and protein prediction

All identified patients had bi-allelic variants in the *Nitrilase 1* gene (*NIT1*, OMIM 604618), located on chromosome 1q23.3, encoding the NIT1 enzyme. Patients were either compound heterozygous for the *NIT1* c.727C>T, p.Arg243Trp variant and the truncating *NIT1* c.198_199del, p.Ala68* variant, or homozygous for the *NIT1* p.Arg243Trp variant (Table 1 and Figure 1A). The *NIT1* p.Arg243Trp variant alters a strongly conserved residue (up to Baker’s yeast) and was predicted to be pathogenic by *in silico* prediction programmes. 3D modelling showed that the p.Arg243Trp variant is located at the interface of the NIT1 monomers comprising the dimeric NIT1 enzyme (Figure 1B). The truncating variant c.198_199del, p.Ala68* introduces a stop codon in exon 3, which is predicted to lead to nonsense mediated decay of the truncated transcript.

**Table 1.**

| Case | F1:II-1 | F1:II-2 | F1:II-3 | F2:I-1 | F3:II-1 | F4:II-1 | F5:II-1 |
| --- | --- | --- | --- | --- | --- | --- | --- |
| **Pedigree** | 1 | 1 | 1 | 2 | 3 | 4 | 5 |
| **Age at last clinical assessment** | †56-60 | 56-60 | †46-50 | 46-50 | 56-60 | 56-60 | 66-70 |
| **Sex** | M | F | M | M | M | M | M |
| ***NIT1* genotype** | *p.Arg243Trp/p.Ala68\** | *p.Arg243Trp/p.Ala68\** | *p.Arg243Trp/p.Ala68\** | *p.Arg243Trp (hom)* | *p.Arg243Trp (hom)* | *p.Arg243Trp (hom)* | *p.Arg243Trp (hom)* |
| **Clinical characteristics (age at onset)** |  |  |  |  |  |  |  |
| Haemorrhagic stroke | + (56-60) | - | + (46-50) | + (46-50) | - | - | + |
| Ischemic stroke | - | - | +<sup>1</sup> | + (36-40) | + | - | NA |
| Tremor | hands (36-40) | hands (36-40) | +<sup>(2)</sup> | - | hands<sup>(2)</sup> | - | NA |
| Dystonia | neck (46-50) | right thumb and index finger<sup>(2)</sup> | +<sup>(2)</sup> | - | - | - | NA |
| Chorea | - | right lower extremity (36-40) | - | left upper extremity<sup>(2)</sup> | - | bilateral upper and lower extremities<sup>(2)</sup> | NA |
| Dysarthria | + (46-50) | +<sup>(2)</sup> | + (36-40) | + | - | +<sup>(2)</sup> | NA |
| Bradykinesia | +/-<sup>(2)</sup> | +<sup>(2)</sup> | - | + | + | + (51-55) | NA |
| Gait disturbance | +/-<sup>(2)</sup> | + (56-60) | - | - | + | + (51-55) | NA |
| Syncope | +/-<sup>(4)</sup> | + (46-50) | NA | + (46-50) | + (56-60) | + (56-60) | NA |
| Fatigue | +<sup>(2)</sup> | +<sup>(2)</sup> | NA | - | - | +<sup>(2)</sup> | NA |
| Psychiatric symptoms | - | depression (41-45) | Psychosis<sup>(2)</sup> | - | - | - | NA |
| Cognitive/memory impairment | + (36-40) | + (51-55) | Unknown | + (36-40) | +<sup>(2)</sup> | +<sup>(2)</sup> | NA |
| MOCA | NA | 10/30 (56-60) | - | 25/30 (46-50) | 18/30 (56-60) | NA | NA |
| **Cardiovascular risk factors\*** | None | HC | HT, HC | HT, HC | None | None |  |
| **Neuroimaging** | MRI + CT | MRI | CT | MRI + CT | MRI | MRI | NA |
| ePVS | + | + | + | + | + | + |  |
| WMH | - | + | Unknown*<br>* | + | + | + |  |
| Lacunes | - | + | Unknown*<br>* | + | + | - |  |
| Microbleeds | – | + | Unknown*<br>* | - | - | + |  |
| Haemorrhagic foci | + | + | Unknown*<br>* | + | - | + |  |
| Intracerebral haemorrhage | + | – | + | + | – | - |  |

<sup>1</sup> Traumatic occlusion of the right internal carotid artery at age 16-20y with left sided hemiparesis with almost full recovery, spontaneous recanalization. Recurrence ischemic stroke right hemisphere at 36-40y. MRA was reported to show an accessory artery originating from the distal carotid artery, suggested to be an arteriovenous malformation.
<sup>2</sup> Age of onset not reported
<sup>3</sup> Feature consistently reported in clinical records, but without further details
<sup>4</sup> Although syncope was not reported, significantly increased sleep tendency with sudden narcolepsy-like sleep attacks was described.
\*Hypertension (HT), hypercholesterolemia (HC), Diabetes Mellitus (DM), smoking (S)
\*\* Could not be assessed on the CT scan made after ICH; MRI was not available for this patient.
Hom=homozygous, NA= not available

**Figure 1.**
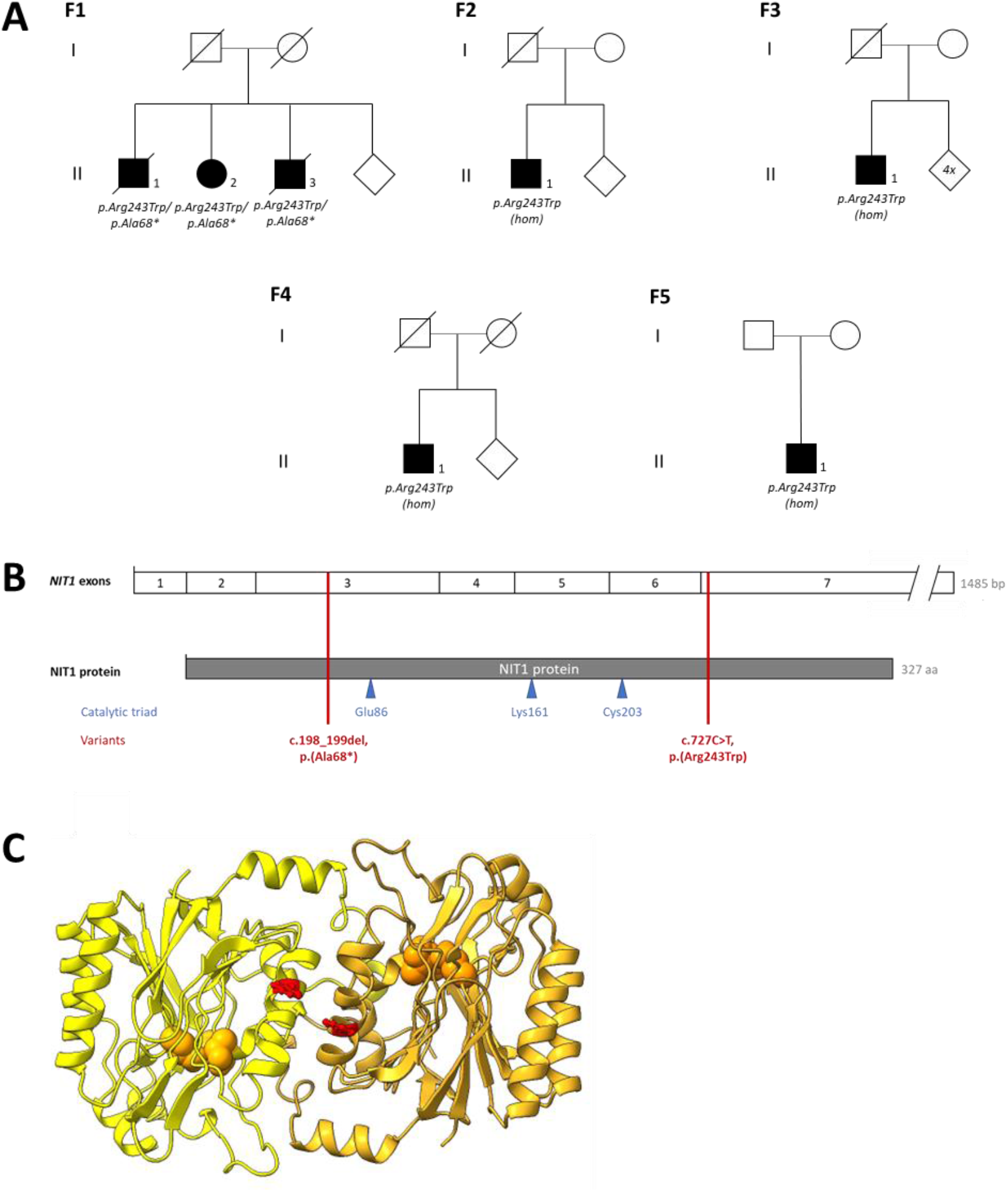
*NIT1* variants detected in five unrelated pedigrees and 3D protein modelling. (A) Simplified pedigrees of the NIT1-small vessel disease patients identified in this study. (B) Schematic representation of the *NIT1* exons and protein, including the location of the amino acids composing the catalytic triad and the *NIT1* variants found in the patients in this study. (C) 3D-modelling of the homodimeric NIT1 enzyme. The p.Arg243Trp substitution (shown in red) is located at the interface of the two NIT1 monomers. Spheric residues indicate the catalytic site of each monomer (Glu86-Lys161-Cys203)

### Clinical and neuroimaging phenotype

Clinical characteristics of patients with bi-allelic *NIT1* variants are summarized in Table 1. Cardinal clinical features included progressive movement disorders such as dystonia, chorea, bradykinesia and tremor, as well as gait disturbance and dysarthria, not as a direct consequence of an evident ischemic of haemorrhagic stroke. Slowly progressive cognitive decline and recurrent syncope was another consistent feature. Two patients died due to massive non-lobar ICH, at age 46-50 and 56-60 years, respectively. A subset of patients also had ischemic strokes. All patients shared a distinct neuroimaging phenotype, with an extremely high burden of enlarged perivascular spaces (ePVS) in both supratentorial and infratentorial brain regions (Figure 2). ePVS were especially prominent in the basal ganglia, thalamus, the mesencephalon, pons and cerebellum, leading to a honeycomb appearance. ePVS were present to a lesser extent in the temporal, occipital and parietal lobes. The burden of white matter hyperintensities varied between patients, even of the same age. Half of the patients had lacunes on MRI. In three patients, hemorrhagic foci in the basal ganglia were seen, located between clusters of ePVS. Neuroimaging (CT) at the time of fatal ICH showed massive haemorrhage in the basal ganglia complex, with intraventricular extension.

**Figure 2.**
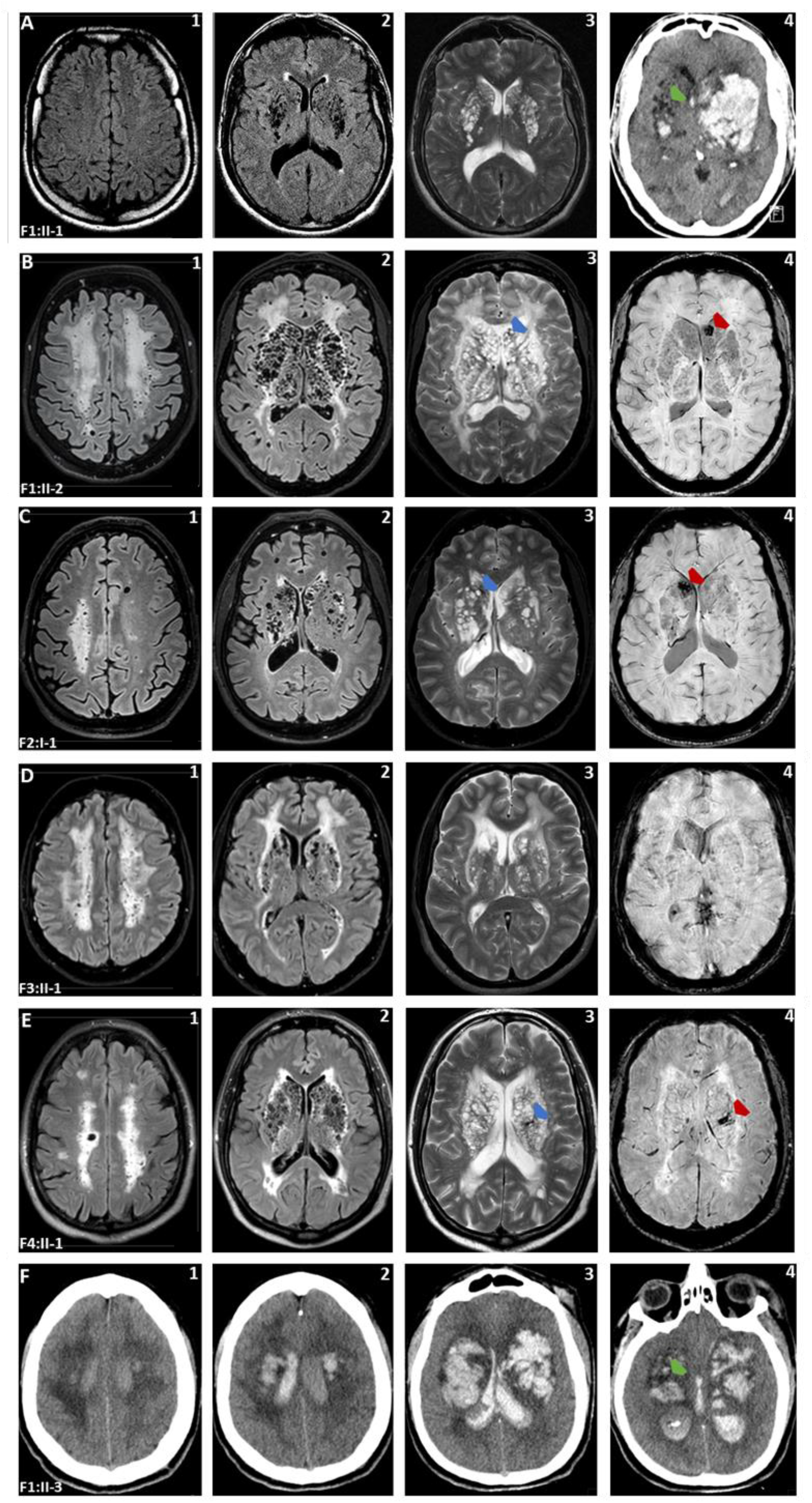
Neuroimaging abnormalities in six patients with NIT1-small vessel disease from four different pedigrees. **A1**,**2-E1**,**2** (T2-FLAIR) White matter hyperintensities and ePVS may be present throughout the brain, including the centrum semiovale. The burden of white matter hyperintensities (WMH) varies strongly, from only limited periventricular capping (**A2**) to extensive confluency of WMH well into the centrum semiovale and the external capsule, mostly symmetrically **(B1**,**2, D1**,**2)** but also unilaterally in one individual after haemorrhagic stroke (**C1**). **A1**,**2-E1**,**2** (T2-FLAIR) and **A3-E3** (T2). A striking état criblé in the basal ganglia with a honeycomb-like appearance due to enlarged perivascular spaces (ePVS) is a commonality in all patients. This is especially well visible on T2-sequences, but focal hypodense lesions indicative of ePVS may also already be observed on CT (green insets on **A4, F4**). Additionally, next to an increased number of ePVS, note the presence of remarkably tumefactive ePVS. Lesions very likely to be lacunes are sometimes interspersed with ePVS, but are in many cases difficult to discern from these tumefactive ePVS. ePVS may be present throughout the brain, from the centrum semiovale **(A1-E1**), to the thalamus and the cerebellum (not pictured). **A4** and **F1-4** (CT). Massive spontaneous bilateral haemorrhage in the basal ganglia with breakthrough into the lateral ventricles. **B4**,**C4**,**E4** (SWI). Susceptibility weighted (SWI) artifacts indicative of hemosiderin deposits are present in the basal ganglia in three patients from three different pedigrees (red insets). These correspond to hypointense lesions located between clusters of ePVS on T2-sequences (blue insets on **B3**,**C3**,**E3**), implicative of haemorrhagic foci. Only a few small focal SWI artifacts suggestive of microbleeds are present in a minority (n=2) of the cases (not pictured).

### Small vessel pathology

The deeper lying perforating arteries and arterioles of the cerebrum, basal ganglia and brain stem showed strongly abnormal vessel wall morphology. There was a thickened media with abundance of hyalin and fibrin and large amorphic deposits which stained positive for Periodic acid–Schiff and toluidine blue (Figure 3A-C). Vessels were negative for Congo Red, Beta Amyloid and NOTCH3 staining. Electron microscopy showed large and extensive granular electron dense deposits in the media and adventitia. Often these deposits were surrounded by fibrillar collagen (Figure 3D,E). Brain venules did not show any vessel wall abnormalities.

**Figure 3.**
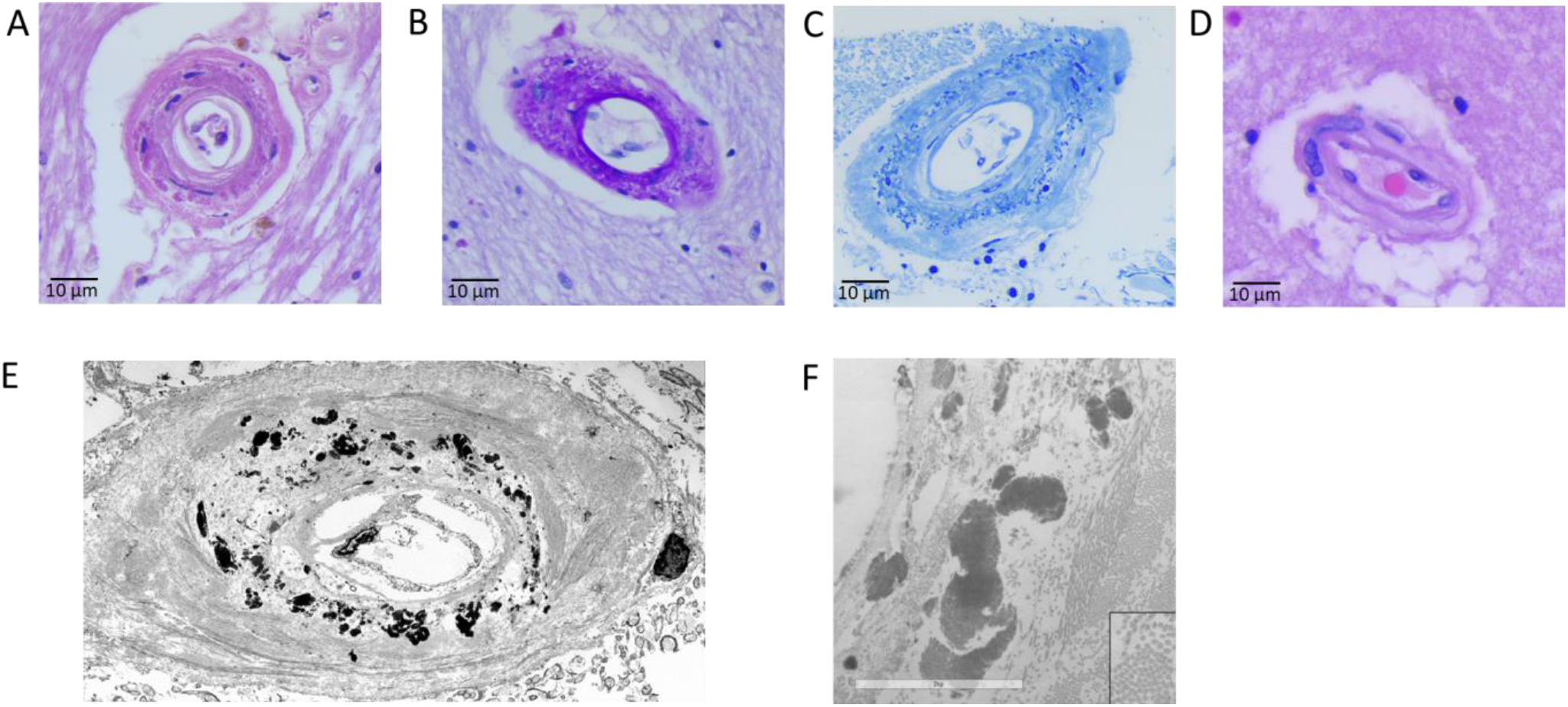
Small vessel wall pathology. (A) Hematoxylin-eosin staining of brain vessels of the two deceased patients from family 1 (II-1 and II-3), showing a thickened media and adventitia with deposits of granular material the outer layers and fibrosis and hyalinization of the more inner layers (B) Periodic acid– Schiff and (C) toluidine blue stain (C) of vessels highlighting the deposits of granular material. (D) Hematoxylin-eosin staining of a brain vessel from a control. (E,F) Electron microscopy of brain vessels showed numerous electron dense deposits in the media and adventitia in both patients. Often these deposits were surrounded by fibrillar collagen. Representative images are shown.

### *NIT1* expression and function in patient tissue

Patient brain tissue showed a lower intensity of NIT1 staining in the grey matter compared to controls. NIT1 staining intensity in the vessels was comparable between patients and controls, except for patient arterioles with a significant loss of smooth muscle cells, which showed reduced NIT1 staining (Figure 4 A,B). Gas chromatography mass spectrometry analysis of urine samples showed increased levels of deaminated glutathione, consistent with loss of NIT1 function (Figure 4 C,D).

**Figure 4.**
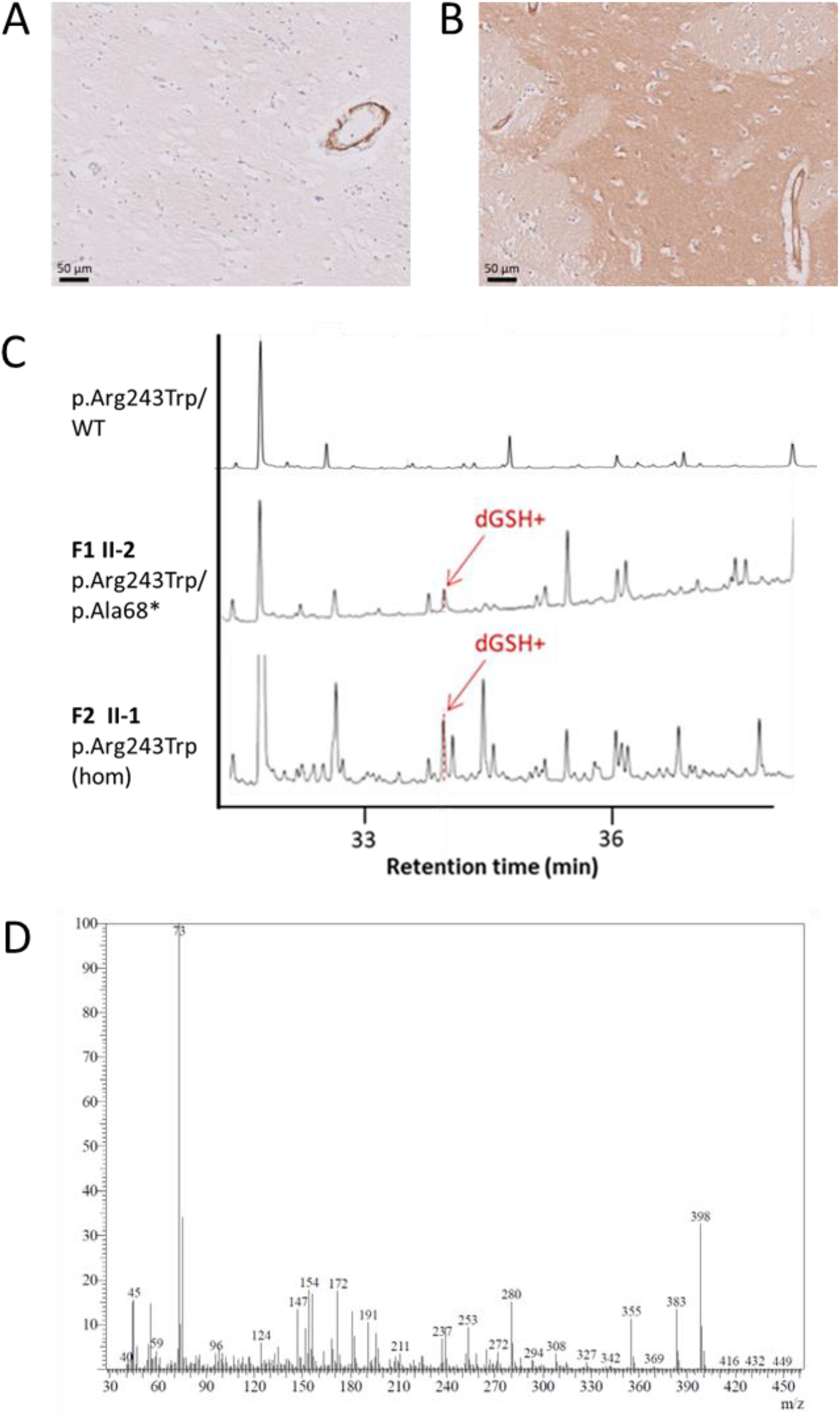
Reduced NIT1 expression and function. (A,B) NIT1 immunohistochemistry on brain tissue (putamen) of NIT1-SVD patients (compound heterozygous for *NIT1* variants p.Arg243Trp and p.Ala68*) showed very weak NIT1 staining of the grey matter in the patients compared to the controls. The smooth muscle cells of the perforating arteries showed a positive NIT1 staining which was comparable between patients and controls. A representative image of one patient (A) and one control (B) is shown. (C) GC-MS analysis of urine of patients F1 II-2 and F2 II-1 showing the presence of the dGSH derivative. Total ion current (TIC) chromatograms were aligned with respect to retention time. The dGSH derivative was not detectable in unaffected heterozygous family members. (D) Mass spectrum of the dGSH derivative peak. The spectrum is essentially identical to the spectrum of purified dGSH.^5^

### *NIT1* variant carrier frequency and estimated *NIT1*-small vessel disease prevalence

The *NIT1* c.727C>T; p.Arg243Trp variant had a carrier frequency of ∼1/2000 in UK Biobank. In gnomAD, the carrier frequency of the p.Arg243Trp variant was 1/4000; p.Arg243Trp was present in the non-Finnish European and African/African American individuals, but not in South and East Asian or Ashkenazi Jewish individuals. p.Arg243Trp carrier frequency was highest in a Dutch population database (GoNL), namely 1/500. The carrier frequency of truncating *NIT1* variants was ∼1/1000 in the various population databases, and truncating *NIT1* variants were seen in individuals from all ethnicities in gnomAD. Based on these frequencies, the minimal estimated prevalence of *NIT1*-small vessel disease in the Netherlands is 1/450,000, and worldwide ∼1/1,800,000. This would mean minimally 40 patients with NIT1-small vessel disease in the Netherlands, and minimally 4500 patients worldwide. The seven Dutch patients were from different regions of the Netherlands, did not come from a genetic isolate and their parents were not known to be consanguineous.

### Heterozygous *NIT1* variant carriers do not have a clinical or neuroimaging phenotype

In UK Biobank, there was no difference in the total neuroimaging burden of white matter hyperintensities or enlarged perivascular spaces between carriers of the *NIT1* variants and controls (Figure 5). None of the *NIT1* variant carriers had neuroimaging evidence of ICH or a medical history of ICH. Ischemic stroke frequency did not differ from controls.

**Figure 5.**
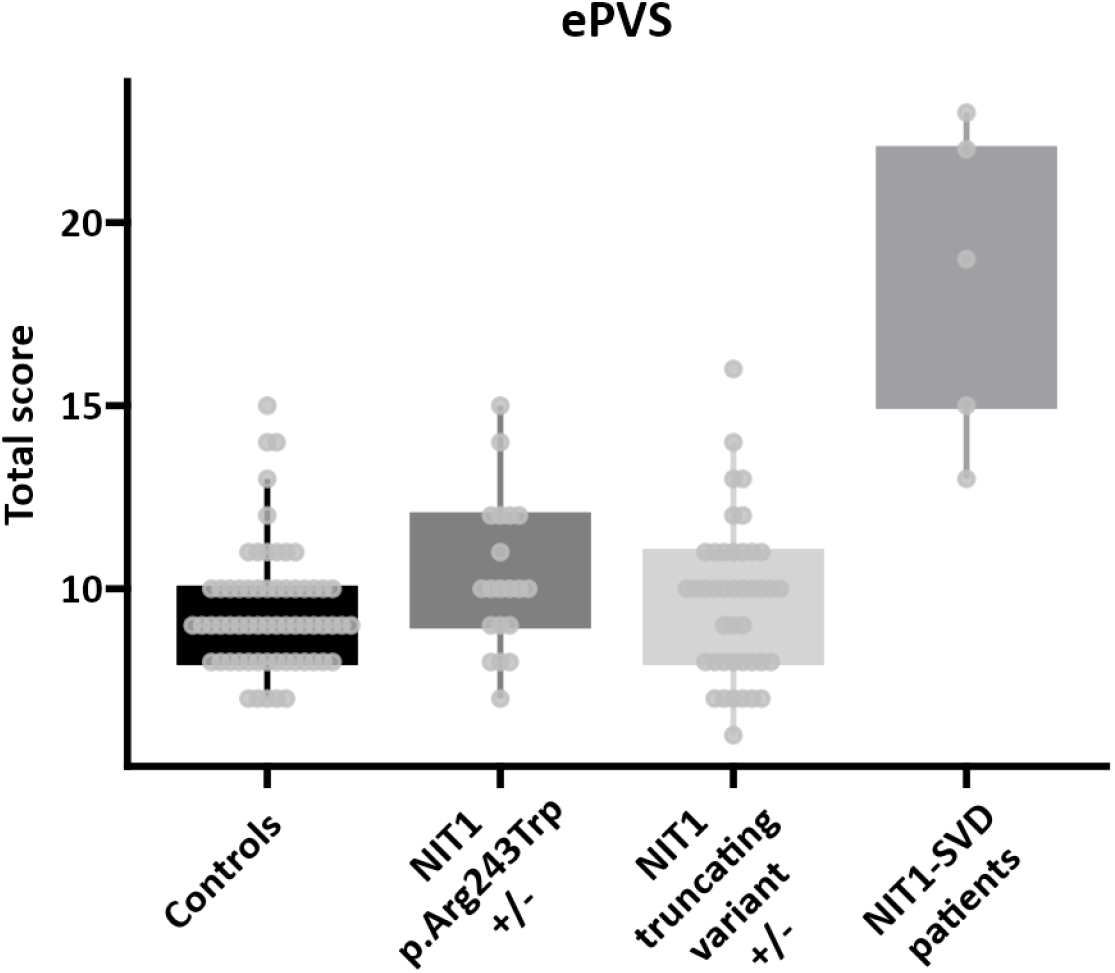
ePVS load in heterozygous *NIT1* variant carriers in UK Biobank compared to controls and NIT1-SVD patients. Individuals from the population with a *NIT1* truncating variant or with the *NIT1* p.Arg243Trp missense variant do not have or enlarged perivascular spaces (B) compared to age and sex-matched controls.

## Discussion

Here, we report a novel genetic cause of intracerebral haemorrhage (ICH), caused by bi-allelic *NIT1* pathogenic variants. We show this to be a novel genetic cerebral small vessel disease (SVD), with extensive electron dense deposits in the media and adventitia of cerebral small arteries and a strongly thickened vessel wall. Patients have striking neuroimaging features, with numerous and strongly enlarged perivascular spaces, resulting in a pathognomonic honeycomb appearance of the basal ganglia-thalamus complex and massive mid-adult onset non-lobar ICH. There are a number of autosomal dominantly inherited causes of ICH, predominantly APP-associated cerebral amyloid angiopathies, CADASIL and COL4A1 and COL4A2-associated SVD. NIT1-small vessel disease (NIT1-SVD) is the first autosomal recessive disorder in which adult-onset ICH is a cardinal feature. The autosomal recessive inheritance can mask the genetic nature of the disorder, as patients often present as apparently sporadic cases. However, the distinctive neuroimaging phenotype enables a spot-diagnosis. After ascertaining the genetic cause in the initial Dutch pedigree, multiple additional cases were diagnosed in our centre based on MRI features and confirmed with *NIT1* sequencing analysis.

Cardinal clinical features of NIT1-SVD are progressive mid-adult onset movement disorders, cognitive decline and fatal non-lobar ICH. Approximately 20 years prior to ICH, patients with NIT1-SVD present with movement disorders such as dystonia, chorea, bradykinesia and tremor, gait disturbance or dysarthria. In addition to disorders of movement, slowly progressive cognitive impairment and recurrent syncope is a key feature. Other reported symptoms include ischemic stroke, extreme fatigue and psychiatric disturbances. The most striking and consistent MRI feature is the honeycomb appearance of the basal ganglia-thalamus complex, due to numerous strongly enlarged perivascular spaces (ePVS). These can be accompanied by haemorrhagic foci seen on susceptibility weighted imaging, located between the clusters of ePVS. Most, but not all patients, have lacunes and extensive confluent periventricular and deep white matter hyperintensities. Two of the patients had microbleeds.

Patients from four unrelated pedigrees were homozygous for the *NIT1* p.Arg243Trp missense variant, whereas three siblings from another pedigree were compound heterozygous for the p.Arg243Trp variant and a truncating variant (p.Ala68*). *NIT1* encodes the highly conserved 36 kD NIT1 protein, which belongs to the nitrilase protein family. *NIT1* is ubiquitously expressed in adult human and mouse tissues and during mouse embryogenesis.^10^ NIT1 is known to act as a metabolite repair enzyme, hydrolysing the deaminated form of the common intracellular antioxidant glutathione (dGSH), thereby yielding alpha-ketoglutarate and cysteinylglycine.^5^ NIT1 has also been shown to act as a tumour-suppressor via activation of the TGFβ–Smad signalling pathway.^10,11^ We confirmed NIT1 loss-of-function in the NIT1-SVD patients by demonstrating increased levels of dGSH in urine, similar to what has been described in *Nit1* knock-out mice.^5^ The currently known functions of NIT1 cannot readily explain the abundant electron dense protein deposits which are present in the (cerebral) small arteries of NIT1-SVD patients. It is highly unlikely that these deposits are composed of dGSH, as dGSH is comprised of only three amino acids and is thus unlikely to form this type of protein aggregates. NIT1 is also an unlikely constituent of the aggregates, as there was no difference in NIT1 staining of affected cerebral arteries of NIT1-SVD patients versus controls. The appearance of the deposits on electron microscopy imaging was strongly reminiscent of the granular osmophilic material seen in CADASIL patients,^12^ but they were much more abundant, larger and more amorphous. Importantly, the vessel wall deposits stained negative for NOTCH3, and also for amyloid. Taken together, this suggests a novel, undiscovered function of NIT1, specifically in vessel wall homeostasis, where loss of NIT1 function leads to aggregation of yet unknown proteins in the (cerebral) vessel wall. Notably, NIT1-SVD patients did not have any other organ dysfunction and no evident predisposition to tumour development.

Bi-allelic *NIT1* loss of function variants have been previously described in a family with psychiatric disturbances and cystic lesions in the basal ganglia.^13^ These cystic lesions are most likely ePVS, as the abnormalities on the single MR image shown in this report are highly reminiscent of the honeycomb appearance of the basal ganglia characteristic for NIT1-SVD.

NIT1-SVD is an adult-onset metabolic genetic small vessel disease. Analogous to other autosomal recessive metabolic disorders, we found that heterozygous carriers of a *NIT1* pathogenic variant did not have a NIT1-SVD clinical or neuroimaging phenotype, and they did not show increased levels of dGSH in urine.

Based on carrier frequencies of the *NIT1* p.Arg243Trp variant and *NIT1* truncating variants in UK Biobank and gnomAD, we calculated that an estimated minimum of 4500 individuals worldwide are affected with NIT1-SVD. In this calculation, we only took into account the p.Arg243Trp variant, which is located at the interface of the two NIT1 monomers that form the dimeric NIT1 enzyme, thereby likely disrupting the formation or stability of the dimer, but other *NIT1* (missense) variants, for example those located in the catalytic triad (Glu86-Lys161-Cys203), may also lead to loss of *NIT1* function, in which case the prevalence of NIT1-SVD would be higher.

The discovery of NIT1-SVD as a novel cause of hereditary intracerebral haemorrhage with a highly distinct neuroimaging phenotype enables correct diagnosis in these patients, often presenting as isolated, seemingly sporadic cases of mid-adult onset movement disorders, followed by non-lobar ICH of unknown cause. After genetic diagnosis, accurate risk estimations and genetic counseling of family members can be provided. Correct diagnosis will also lead to personalized medical management, for example by being reticent with regard to the prescription of antiplatelets after lacunar stroke, or the use of anticoagulants, as these may elicit or aggravate ICH. The fact that NIT1-SVD is a metabolic disorder with loss of NIT1 enzymatic function, may provide avenues for future enzyme replacement therapy.

In conclusion, we describe a novel cause of genetic cerebral small vessel disease characterized by movement disorders, progressive cognitive decline, a highly distinct neuroimaging phenotype and non-lobar ICH, caused by bi-allelic *NIT1* loss-of-function variants.

## Data Availability

All data produced in the present study are available upon reasonable request to the authors

## Acknowledgements

The authors thank the patients and their family members for participating in the study, and M. van der Knaap, T. van Osch and E. Oudeman for their contribution to patient identification and clinical characterization. This study was performed using the UK Biobank resource, under Application Number 74162.

